# Estimates of cases and hospitalizations averted by COVID-19 case investigation and contact tracing in 14 health jurisdictions in the United States

**DOI:** 10.1101/2021.05.27.21257931

**Authors:** Seonghye Jeon, Gabriel Rainisch, R. Ryan Lash, Patrick K. Moonan, John E. Oeltmann, Bradford Greening, Bishwa B. Adhikari, Contact Tracing Impact Group, Martin I. Meltzer

## Abstract

**Context:** The implementation of case investigation and contact tracing (CICT) for controlling COVID-19 (caused by SARS-Cov-2 virus) has proven challenging due to varying levels of public acceptance and initially constrained resources, especially enough trained staff. Evaluating the impacts of CICT will aid efforts to improve such programs.

**Objectives:** Estimate the number of COVID-19 cases and hospitalizations averted by CICT and identify CICT processes that could improve overall effectiveness.

**Design:** We used data on proportion of cases interviewed, contacts notified or monitored, and days from testing to contact notification from 14 jurisdictions to model the impact of CICT on cumulative cases counts and hospitalizations over a 60-day period. Using the Centers for Disease Control and Prevention (CDC)’s COVIDTracer tool, we estimated a range of impacts by assuming either contacts would quarantine only if monitored or would do so upon notification of potential exposure. We also varied the observed program metrics to assess their relative influence.

**Results:** Performance by jurisdictions varied widely. Jurisdictions isolated between 12 and 86% of cases (including contacts which became cases) within 6 to 10 days after exposure-and-infection. We estimated that CICT-related reductions in transmission ranged from 0.4% to 32%. For every 100 cases prevented by nonpharmaceutical interventions, CICT averted between 4 and 97 additional cases. Reducing time to case isolation by one day increased averted case estimates by up to 15 percentage points. Increasing the proportion of cases interviewed or contacts notified by 20 percentage points each resulted in at most 3 or 6 percentage point improvements in averted cases.

**Conclusions:** We estimated that case investigation and contact tracing reduced the number of COVID-19 cases and hospitalizations among all jurisdictions studied. Reducing time to isolation produced the greatest improvements in impact of CICT.

## INTRODUCTION

Case investigation and contact tracing (CICT) is a core public health activity routinely used to reduce transmission of infectious diseases. However, COVID-19 CICT programs have found it difficult to keep pace with exponential growth in caseloads. As early as April 2020, reports surfaced that the United States public health workforce was insufficient to keep pace with new, COVID-19 pandemic infections.^1^ Additionally, public acceptance and participation in CICT programs has been low in some jurisdictions.^2-4^ Health departments were also challenged, particularly in the early phases of the pandemic, to balance financial and human resource allocations for these programs with other community-wide interventions and policies.^3, 5^ These difficulties, created considerable variation in how COVID-19 CICT was implemented across the United States.^3^

Several modeling studies suggest that CICT for COVID-19 can be effective; however, these were based on simulation data rather than data collected from health departments.^6, 7^ While a few studies have reported on COVID-CICT program performance in the United States, only two studies systematically evaluated programs from multiple jurisdictions.^2, 4, 8, 9^ Lash *et al*.^2^ reported that while CICT programs in 14 jurisdictions could successfully identify and interview cases, only 1 in 3 cases reported any contacts to be notified. Spencer *et al*.^9^ reported that higher caseloads per contact tracer were correlated with longer times to contact notifications. Neither study, however, estimated the impact of the reported levels of effectiveness in terms of cases averted.

In this study we use metrics from CICT (*e*.*g*., proportion of cases interviewed, contacts notified or monitored, and days from index case testing to contact notification) from the 14 jurisdictions assessed by Lash *et al*.^2^ to estimate the number of potential cases and hospitalizations averted. We also varied the program metrics to assess their relative influence and identify how public health officials might improve the effectiveness of CICT.

## METHODS

### Overview

We used the Centers for Disease Control and Prevention (CDC)’s COVIDTracer Advanced modeling tool^10^ to estimate cases and hospitalizations averted due to CICT programs in 14 health departments (HDs) from 11 states (including six counties, two health districts [*i*.*e*., several adjacent counties], 5 entire states, and one tribal nation) over a 60-day observation period. The 60-day observation periods occurred in the time period spanning from approximately June through October 2020 (before COVID-19 vaccines were approved), and were based on locations’ availability to participate in the Lash *et al*. evaluation. COVIDTracer Advanced is a spreadsheet-based modeling tool, estimating the spread of COVID-19 and impact of interventions (including CICT strategies), in a user-defined population. All locations included in this study employed, concurrent with their CICT efforts, other nonpharmaceutical interventions (NPIs), such as facemask policies, restrictions on large gatherings, and school and business closures. We used COVIDTracer Advanced to proportionately attribute reductions in transmission to either CICT or all other NPIs (details below).

### Assessing Net Impact

We assessed the net impact of CICT in each jurisdiction by calculating, the minimum and maximum cases and hospitalizations averted. We also calculated the percent of cases, or hospitalizations, averted due to CICT, as a percent of total cases averted if only NPIs were implemented. Essentially, for every 100 cases (or hospitalizations) prevented by NPIs, we calculated the additional cases (and hospitalizations) averted due to CICT. These percentages allowed for comparisons of CICT impact between different sized jurisdictions.

### Defining Case Investigation and Contact Tracing Effectiveness

We defined CICT effectiveness as a combination of the proportion of cases (including contacts which became cases) that were effectively isolated, and the time taken from exposure-and-infection to effective isolation. We defined effective isolation (for confirmed cases) and effective quarantine (for contacts of confirmed cases) as being placed, or going into isolation or quarantine such that onward transmission of COVID-19 is essentially stopped. The duration of quarantine and isolation is described in CDC’s interim guidance.^11^ We used data observed by Lash *et al*.^*2*^ at each location (Table 1). Observations from each location provided the time from specimen collection to when the public health department interviewed the cases, and subsequently notified contacts. As there were no data recorded on when a contact was exposed and potentially infected, we assumed that on average, there were 5 days from when a case was exposed-and-infected to when they became symptomatic,^12, 13^ and that cases were tested (*i*.*e*., specimen collected) the day following symptom onset. We further assumed that interviewed cases and contacts isolated and quarantined themselves the day after being interviewed by the program. We conducted sensitivity analyses on the number of days from exposure-and-infection to quarantine and isolation. Detailed descriptions of the estimated time from exposure-and-infection to isolation and quarantine are provided in the Supplementary Content.

**Table 1.** Observed Case Investigation and Contact Tracing Performance Measures and Summarized Program Effectiveness for Selected Locations^*^

|  | **Location 1** | **Location 2** | **Location 3** | **Location 4** |
| --- | --- | --- | --- | --- |
| **Population†** | Category B | Category A | Category C | Category A |
| **COVID-19 Incidence<sup>a</sup>** | <1/100k pop | 6/100k pop | 12/100k pop | 26/100k pop |
| **Performance Measures (Observed<sup>b</sup>)** |  |  |  |  |
| **Community Receptivity Metrics** |  |  |  |  |
| % of cases interviewed | 99% | 83% | 91% | 33% |
| % of interviewed cases named contacts | 87% | 87% | 44% | 21% |
| **Program Logistics Metrics** |  |  |  |  |
| % of all contacts identified<sup>c</sup> | 86% | 72% | 40% | 7% |
| % of identified contacts notified | 95% | 61% | 85% | 54% |
| % of identified contacts monitored | 19% | 56% | 0% | 48% |
| Timing of test results notification (days post specimen collection)<sup>d</sup> | 2 days | 3 days | 2 days | 5 days |
| Timing of contact notification (days post specimen collection)<sup>e</sup> | 2 days | 5 days | 7 days | 7 days |
| **Summary Effectiveness Measures (Calculated<sup>f</sup>)** |  |  |  |  |
| **% of cases and contacts isolated<sup>g</sup>** | 39.5 – 86.0% | 52.7 – 55.0% | 26.1 – 50.5% | 11.8 – 12.1% |
| **Days from infection to contact isolation<sup>h</sup>** | 6 days | 8 days | 9 days | 10 days |

\* Equivalent data for all 14 jurisdictions available in Supplemental Digital Content 1 Table A4.
† To preserve anonymity, location populations are categorized by population size as follows: Category A > 1 million; Category B >500 000 to ≤ 1 million; Category C: ≤ 500 000.
<sup>a</sup> Mean daily incidence for the 60 days of the observation period.
<sup>b</sup> From Lash *et al.*<sup>2</sup>
<sup>c</sup> The number of named contacts divided by the expected number of contacts per case (Supplemental Content; Equations for Case Investigation and Contact Tracing Effectiveness).
<sup>d</sup> This is the reported median days from specimen collection to positive test result reported to health departments.
<sup>e</sup> This is the reported median days from specimen collection to contact notification.
<sup>f</sup> These values were used as inputs in COVIDTracer Advanced. They were calculated from the Observed Performance Measures. See Supplemental Content, Equations for Case Investigation and Contact Tracing Effectiveness section.
<sup>g</sup> Low value is the minimum estimate of CICT effectiveness, and assumes monitoring is required for effective isolation of contacts. High, or maximum, value assumes contact notifications are sufficient for behavior changes in notified contacts such that there is effective isolation of contacts.
<sup>h</sup> The average length of time from exposure-and-infection to isolation and quarantine of cases, and contacts who became cases.

Note, a portion of cases reported to the health department were unable to be reached for interview by CICT programs, and even when reached, some cases chose not to complete the interview or did not name potential contacts (Table 1 and Supplemental Digital Content 1 Table A4).^2, 9^ Finally, we assumed that our calculated program effectiveness values were constant (*i*.*e*., not time-varying) over our 60-day observation periods.

### Estimating Cases Averted

As mentioned earlier, we used COVIDTracer Advanced to proportionately attribute reductions in transmission due to either CICT or all other NPIs. First, we entered epidemiological data that defines the transmission of COVID-19 into COVIDTracer Advanced (*e*.*g*., daily risk of onward transmission since exposure-and-infection, persons infected per infectious person, % cases that are asymptomatic; Supplemental Digital Content 1 Tables A1 and A2). Then, to define effectiveness of CICT in each jurisdiction, we entered into COVIDTracer Advanced the estimated days from infection to isolation and the days from infection to contact quarantine (Table 1, Supplemental Digital Content 1 Table A4). We also entered each jurisdiction’s population and daily COVID-19 cases^14^ (Supplemental Digital Content 1 Table A4). For each jurisdiction we plotted the cumulative total reported cases for the relevant time period. We then entered an initial estimate of the level of reduction in transmission due to NPIs, thus causing COVIDTracer Advanced to produce an initial plot of cumulative cases over the 60-day period after accounting for reduction in transmission due to both CICT and NPIs. This plot was then compared to the plot of actual reported cases. We then adjusted our initial estimate of the effectiveness of NPIs until the resultant plot of the cumulative cases produced by COVIDTracer Advanced closely matched the plot of actual cases (*i*.*e*., we “fitted the plot” – Supplemental Digital Content 1 Figure A1). After estimating the effectiveness of NPIs we then “ switched off” CICT-attributed effectiveness (by setting its effectiveness to zero) in COVIDTracer Advanced while maintaining the estimated NPI effectiveness. This simulated what would have happened if CICT had not occurred.

### Cases Averted: Minimums and Maximums

For each jurisdiction we calculated a minimum and a maximum of all cases effectively isolated due to CICT (Supplementary Material, *Equations 1–2*). To calculate a maximum, we assumed that all notified contacts effectively quarantined after notification of their exposure (regardless of any monitoring). That is, we assumed that just the notification of exposure would be sufficient to change the contact’s behavior and cause the contact to quarantine. To calculate a minimum, we assumed that effective quarantine was only achieved if a contact agreed to quarantine and be actively monitored for adherence to such isolation requirements (*i*.*e*., follow-up phone, text, or email inquiries after the initial notification from public health authorities).

### Estimating Hospitalizations Averted

The number of averted hospitalizations was calculated in COVIDTracer Advanced by multiplying the estimated number of averted cases by age-stratified infection-to-hospitalization rates. To simplify and maintain anonymity of locations, we used the COVIDTracer Advanced age-based default values for risks of hospitalizations for all 14 jurisdictions (Supplementary Content, Table A3).

### Case Investigation and Contact Tracing Scenarios

Although we analyzed the impact of CICT in all 14 locations, we selected four locations to highlight the range of health impacts associated with CICT implementation and the characteristics that influence program effectiveness. Our selected locations illustrate different combinations of COVID-19 burden, community receptivity to CICT (illustrated by factors such as willingness of cases to name contacts, and contacts’ willingness to agree to be monitored whilst in quarantine), and time from patient being tested to when the CICT program interviewed cases and contacts (Table 1).

Among all 14 jurisdictions, Location 1 had the lowest COVID-19 incidence (<1 daily care per 100 000 population), the greatest community receptivity to CICT, and matched several jurisdictions able to notify contacts most quickly. Location 4 is characterized by values on the other extreme: the 4^th^ highest daily incidence (26 daily cases per 100 000 population), lowest receptivity to CICT, and longest time to notify contacts among all 14 jurisdictions. Locations 2 and 3 are characterized by values nearer to the medians of each measure. Supplemental Digital Content 1 Table A4 contains data similar to that in Tables 1 and 2 for all 14 locations.

**TABLE 2.** Estimated impacts of case investigation and contact tracing (CICT), and nonpharmaceutical interventions (NPIs), by location^a^ over 60-day period after contact tracing evaluations initiated

|  | **Location 1** | **Location 2** | **Location 3** | **Location 4** |
| --- | --- | --- | --- | --- |
| **Transmission Fraction** |  |  |  |  |
| Reduction from CICT | 8.6 – 26.2% | 5.0 – 5.2% | 1.4 – 2.7% | 0.4 – 0.4% |
| Reduction from NPIs<sup>b</sup> | 54.6 – 36.6% | 57.6 – 57.3% | 63.5 – 62.0% | 61.0 – 61.0% |
| Remaining Transmission<br>(100% minus above values) | 36.8 – 37.2% | 37.4 – 37.5% | 35.1 – 35.3% | 38.6 – 38.6% |
| **Additional Cases Averted by**<br>**CICT (%<sup>c</sup>), 60 days** | 651 – 9 480<br>(67.1 – 96.8%) | 12 598 – 13 568<br>(47.1 – 48.8%) | 344 – 768<br>(15.4 – 28.8%) | 859 – 882<br>(4.4 – 4.5%) |
| **Additional Hospitalizations**<br>**Averted by CICT (%<sup>c</sup>), 60**<br>**days** | 16 – 233<br>(67.1 – 96.8%) | 310 – 333<br>(47.1 – 48.8%) | 8 – 19<br>(15.4 – 28.8%) | 21 – 22<br>(4.4 – 4.5%) |

**Notes**
<sup>a</sup>CICT implemented per scenarios in Table 1 and effects were assumed constant over 60 days.
<sup>b</sup>Nonpharmaceutical interventions including masks use, social distancing, school and restaurant closures, etc. Low NPI effectiveness values were generated with the fitting process when CICT effectiveness was high; similarly, high NPI effectiveness values were generated when CICT effectiveness was low.
<sup>c</sup>Percent calculated as (Total Cases Averted when both CICT and NPIs implemented - Total Cases Averted when only NPIs implemented) / Total Cases Averted when both CICT and NPIs implemented). Essentially, for every 100 cases prevented by nonpharmaceutical interventions, CICT averted between 4.4 and 96.8 additional cases

### Sensitivity Analyses

For the 4 selected locations (Table 1), we evaluated the relative importance of three process measures to determine which had the most influence on the impact of CICT. The three process measures that we varied were: 1) the proportion of cases interviewed, varied by 20 percentage points higher and lower; 2) the percentage of contacts notified or monitored, also varied by 20 percentage points higher and lower; and 3) the time (in days) needed to isolate cases, varied by one to three days, faster and slower. When a location’s baseline proportion of cases interviewed or contacts notified or monitored was above 80% (or below 20%), we capped the varied proportion at a maximum of 100%, or a minimum of 0%.

We also examined the effects of adjusting the reported cases for under-reporting and under-detection of cases. It is estimated that up to 40% of COVID-19 cases never develop any symptoms (*i*.*e*., asymptomatic cases) and are likely not detected or reported.^15^ In addition, CDC’s seroprevalence surveys found that cases may be under-reported and under-detected by a factor of 2-5 in the US, with large variabilities across jurisdictions.^16^ Therefore, we performed a sensitivity analysis by increasing the reported cases by a factor of two and then refitting the epidemic curves to estimate impacts of CICT.

## RESULTS

### Case Investigation and Contact Tracing Effectiveness

CICT metrics varied across the 14 locations, from 12% of cases and contacts isolated or quarantined ten days post infection at Location 4, to 86% of cases and contacts isolated or quarantined six days post infection at Location 1 (Tables 1 and A4). Assuming a 3-day pre-infectious period (Table A1), the days before isolation or quarantine equate to a range of three to seven days of potential transmission before infected contacts were quarantined in Locations 1 and 4, respectively.

The differences between a location’s minimum and maximum estimates of cases averted were greatest when the difference between the percentage of contacts notified and actively monitored were also large. For example, in Locations 1 and 3, the percent of contacts monitored was less than a quarter of the contacts notified (Table 1). This caused the maximum cases averted estimated for these two locations to exceed the minimum by at least twice the minimum estimate itself (Table 2).

### Cases and Hospitalizations Averted

In Location 1, over a 60 day period, the CICT program averted 67 – 97% of additional cases and hospitalizations, measured as percentages of cases and hospitalizations averted if only NPIs were implemented, (Table 2 and Figure 1).

**FIGURE 1.**
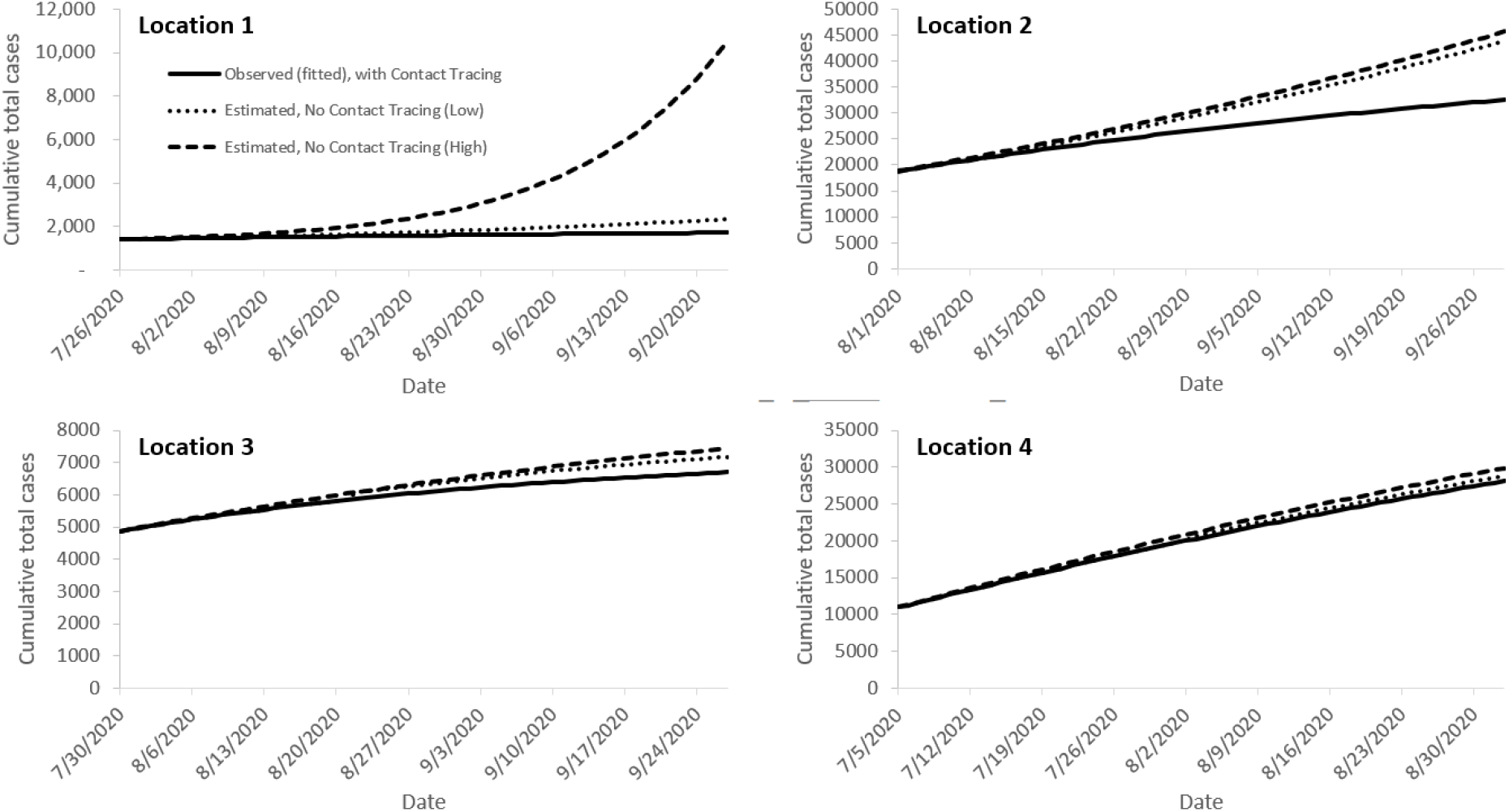
Epidemic curves fitted to observed case counts with case investigation and contact tracing programs, and estimated cases illustrating what might have occurred had the programs not been implemented **Notes**. Solid lines are the epidemic curves at four evaluation locations (1, 2, 3, and 4) fitted to their observed cumulative case counts with both case investigation and contact tracing (CICT) and nonpharmaceutical interventions (NPIs) implemented (Figure 4A). Dashed and dotted lines are the estimated curves illustrating the maximum (dashed) and minimum (dotted) cumulative total cases that might have occurred at these locations if CICT had not been implemented, and only NPIs were implemented, during the 60-day period. The differences between the solid and dashed or dotted lines show the benefits of CICT under minimum and maximum assumptions of program effectiveness, with greater divergence between the solid and broken lines indicating greater impact. All results assume that the effects of CICT and NPIs were constant over the 60 days shown.

This equated to 651 – 9 480 CICT-averted cases and 16 – 233 CICT-averted hospitalizations. In contrast, at Location 4, less than 1% of additional cases and hospitalizations were averted by CICT over 60 days compared to a scenario where only NPIs were implemented (Table 2 and Figure 1). However, due to Location 4’s sizeable population (Table 1), the absolute effects of CICT on transmission were not trivial, with approximately 900 cases averted during the 60-day observation period (Table 2). At Location 2, CICT averted 47 – 49% of cases and hospitalizations (as percentage of cases and hospitalizations averted if only NPIs were implemented), and at Location 3, 15 – 29% were averted by CICT (Table 2).

### Results: Sensitivity Analyses

The time from infection of cases to their isolation and contacts’ quarantine had the most substantial impact on the number of cases and hospitalizations averted in all four locations (Figure 2 and Supplementary Content Table A5a–b, Figure A3). Our results suggest if locations had reduced their baseline times from case exposures to isolation by one day, they would have increased the percentage of cases and hospitalizations averted (compared to cases averted if only NPIs had been implemented) by as much as 15 percentage points (Location 2). Increasing the percent of cases interviewed or contacts notified by 20 percentage points would have resulted in at most 3 or 6 percentage points increases in CICT-averted cases and hospitalizations (Figure 2).

**FIGURE 2.**
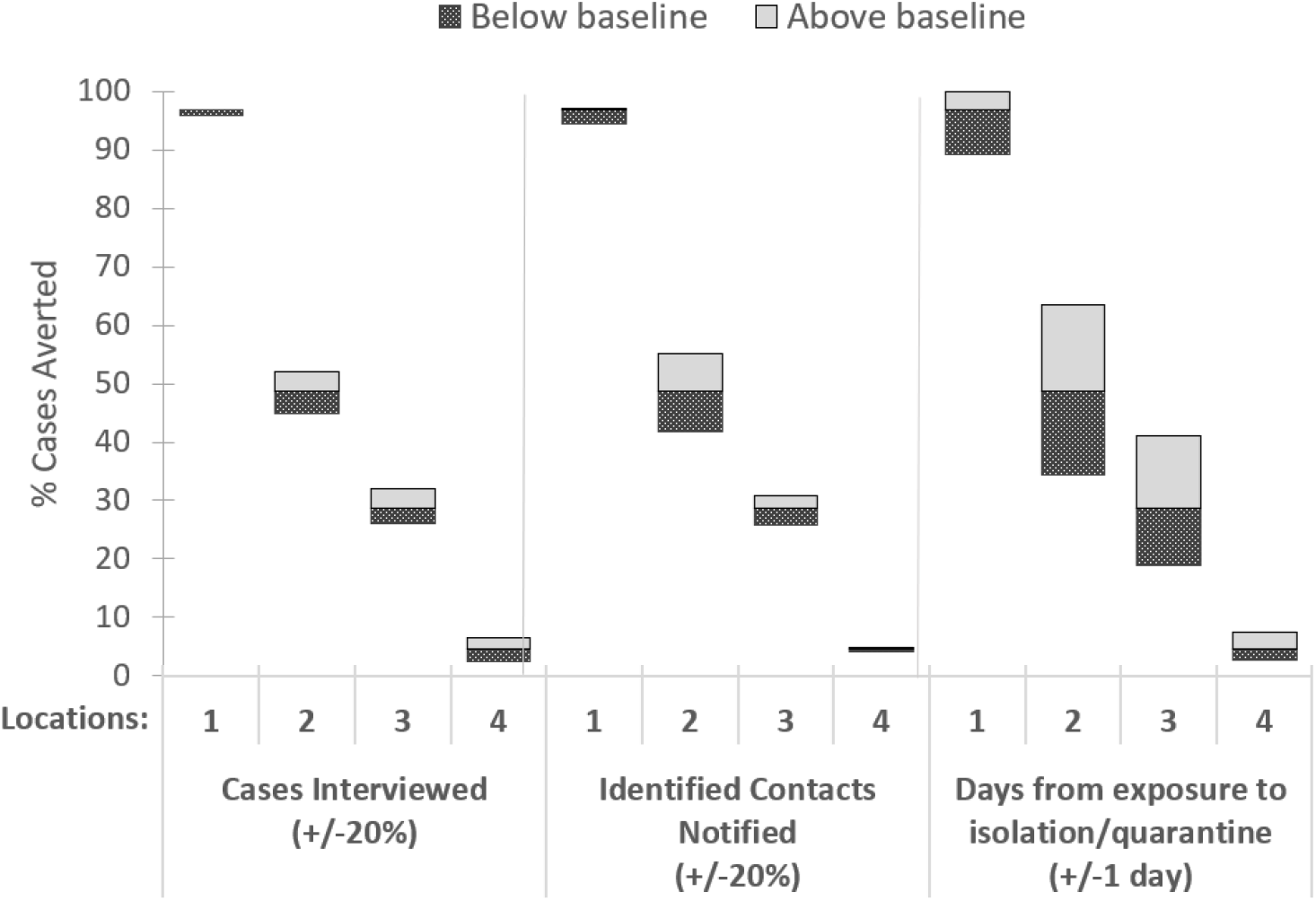
Effects of improvements and constraints to case investigation and contact tracing performance measures compared to the baseline percent cases averted by the programs* **\* Percent cases averted by cases investigated and contact tracing (CICT) calculated as percentage of total cases averted if only nonpharmaceutical interventions (NPIs) were implemented.** **Notes**. Results shown assume notification is a sufficient trigger for contacts to quarantine (maximum effectiveness CICT). Results for our minimum CICT effectiveness scenario (*i*.*e*., monitoring contacts is necessary for effective quarantine) are provided in Figure A3. Baseline results are shown in Table 2. Cases Interviewed and Contacts Notified were capped at 100% and 0% when the baseline percentage interviewed was greater than 80% or less than 20%. See Supplementary Content Table A5a-b for more detailed results.

We found that locations with a longer baseline number of days between case exposures and their isolation experienced greater impact from reducing this time. For example, Location 2 could have increased the proportion of cases averted by 15 percentage points simply by speeding up isolation from 9 to 8 days. In contrast, if Location 1 reduced its time to case isolation from 6 to 5 days, cases averted would have improved by only 0.3% (Supplemental Digital Content 1 Table A5a).

For the sensitivity analysis examining the impact of under-reported and under-detected cases (where we increased reported cases by a factor of two), we found that such adjustment leads to increased estimates of cases averted due to case investigation and contact tracing (Supplemental Digital Content 1 Table A7). However, the fractional attribution of the reduction in transmission due to case investigation and contact tracing remained similar to our original estimates (Table 2).

## DISCUSSION AND CONCLUSIONS

Our modeled estimates of cases and hospitalizations averted illustrate that CICT can be effective in containing the impact of COVID-19. Impacts due to CICT varied widely across the locations studied, averting 4% of cases and hospitalizations in one locale to 97% in another from among the estimated cases and hospitalizations in a scenario where only NPIs were implemented. These estimated reductions were also substantial on an absolute basis, ranging from 859 averted cases (0.1% of Location 4’s population) to 9 480 (1.5% of Location 1’s population) over a 60-day period. Such sizeable estimates of averted cases are partially due to the success of CICT programs at suppressing the exponential growth in cases associated with uncontrolled transmission (2.5 new infections per case without interventions and almost all the population susceptible to infection [Table A2]), compounded over approximately 10 generations of infection that occur in our 60-day observation period. For example, at Location 1, where our estimates of CICT impacts were greatest, our low estimate of the effects of NPIs on transmission (a 36.6% reduction) caused transmission to drop from 2.5 infections per infectious case to 1.6. When we add our estimated effect of CICT to the transmission reductions already afforded by NPIs (a further 26.2% reduction in transmission), the effective number of new infections per infectious case drops to below 1 (0.93). An epidemic is waning any time that the number of new infections per infectious case drops below 1.0. We estimated large percentage reductions in cases (90%+) at locations where the combined effects of NPIs and CICT approached (or dropped below) 1 new infection generated per case (Locations 1, 5, and 6).

Our modeled estimates indicate that reducing the time from notification of a positive test result and the subsequent isolation and quarantine of cases and contacts (even by just 1 day) provides the biggest gain in improving the impact of CICT. We found this to be true even when compared to the potential impact of a 20% increase in percentage of cases interviewed or contacts notified and monitored for compliance with quarantine recommendations. For example, even if locations 2 and 3 had interviewed 100% of their cases and notified 100% of the identified contacts, they would not have achieved the same gains in cases and hospitalizations averted as if they had reduced the time to case isolation by a single day. Our results were consistent with previous studies that found that minimizing testing delay or onset-to-isolation delay had the largest impact on reducing onward transmissions.^6, 17, 18^ The benefits from doing so are most pronounced (*i*.*e*., will yield the greatest impact on cases averted) among jurisdictions with longer delays (8 days or longer after case exposure-and-infection) to notify and isolate contacts.

It should be noted that we do not attribute to any single factor the potential impacts of reductions in time from exposure to isolation and quarantine. Our estimates of additional averted cases could have been achieved from any combination of many factors, such as: earlier testing of cases, less time between patient testing and test results reported to the health department, shorter time from test results received to contact notification, or from contacts choosing to quarantine more quickly. While these and other process metrics can be improved by increasing the number of case investigators and contact tracers or with automated systems and technology upgrades, options exist that may not require many additional resources. For example, health departments can prioritize interviewing cases or notifying contacts of COVID-19 cases most recently tested, or those experiencing fewer days of symptomatic illness. Additionally, when contact tracers’ caseloads are high, devoting less time to cases and contacts that are unreachable or do not respond to messages may reduce effective time from exposure-and-infection to isolation and quarantine.

This study has several strengths. First, this work can be replicated by other jurisdictions that are interested in estimating direct health impact of CICT activities. The tool we used, COVIDTracer Advanced, is publicly available and designed for use by practicing public health officials, although its use for this analysis required advanced familiarity of features.^10^ In addition, by aggregating and presenting systematically collected data from multiple anonymized jurisdictions, we were able to show the range of potential effects, from health departments experiencing resource constraints to those with ample support.

This study also has several limitations. First, we did not directly measure the proportions of cases that effectively isolated and contacts that correctly quarantined. In the absence of data suggesting otherwise, we assumed cases and contacts fully comply with isolation and quarantine guidance they received via their interactions with health departments. As such, our results may overestimate the absolute impact of CICT (and underestimate the effectiveness of NPIs), with our lower effectiveness results likely approximating real impacts more closely. Our conclusions, however, regarding the relative influence of CICT measures are not affected by this limitation.

Additionally, our maximum estimates of effectiveness offer insight into the potential benefits from high public compliance with quarantine guidance. Nevertheless, there is obvious need for additional research to improve our understanding of the actual efficacy of isolation and quarantine, based on public compliance, the role of health departments in motivating such, as well as practical limitations (*e*.*g*., inadequate sick leave, caregiving responsibilities). Lastly, we do not know whether Exposure Notification (EN) Apps (*i*.*e*., smartphone-enabled contact tracing apps) were utilized by locations during the observation period we analyzed, and if so, how their use would have affected our results. A second limitation is we assumed that the effectiveness of CICT and NPIs remained constant over a 60-day period. Since we were able to approximate the observed cumulative epidemic curves for most locations we studied, this assumption appears reasonable for our observation period (Figure A1). Another limitation is that, in using the COVIDTracer Advanced age-based default values for risks of hospitalizations for all 14 jurisdictions, we do not account for the potential that age-based admissions of patients with COVID-19 infections may differ from one location to another. As such, we may over- or under-estimate the hospitalizations prevented. We believe, however, that it is unlikely that incorporating location-specific precision on hospitalizations would yield any different policy choices. Finally, we do not quantify or account for the resources and effort necessary to achieve the gains possible by focusing on notification speed over increasing the proportions of cases interviewed and contacts notified.

Our analysis combined primary implementation data with modeling to assess the health impact of COVID-19 case investigation and contact tracing in the United States. While we illustrated that CICT can be effective at reducing onward transmission in some jurisdictions, other jurisdictions saw limited effectiveness. The exact reasons for, and the relative importance of the potential factors causing such reduced levels of impact remain to be investigated. This work provides public health decision makers with an understanding of the benefits that CICT programs can produce, as well as ideas for prioritizing potential improvements of such programs.

### Implications for Policy and Practice

- Case investigation and contact tracing can be effective in preventing COVID-19 infections and hospitalizations.
- Greatest improvements in the impact of COVID-19 case investigation and contact tracing can be achieved by reducing the time to notify cases and contacts (even by just one day, presuming subsequent isolation and quarantine). The benefits from reducing time-to-notification will be most pronounced among jurisdictions with longer delays between when patients provide a sample for testing and when cases and contacts are notified and requested to isolate or quarantine.
- Jurisdictions can potentially improve the speed of notification without additional resources by (1) prioritizing cases and their contacts that are most recently tested (or those experiencing fewer days of symptomatic illness); (2) devoting less time to cases and contacts that are unreachable or do not respond to messages; (3) limiting the duration of calls (*e*.*g*., using less questions or a shorter script) during periods of high caseloads.

## Supporting information

Supplemental Digital Content

## Data Availability

Data is available upon request

## Acknowledgements

Chinle Service Unit, Indian Health Service, Chinle, AZ; Epidemic Intelligence Service, CDC, Atlanta, GA; Gwinnett, Newton, Rockdale Counties Health Departments, Lawrenceville, GA; Mecklenburg County Public Health, Charlotte, NC; Nebraska Department of Health and Human Services, Lincoln, NE; North Carolina Department of Health and Human Services, Raleigh, NC; New Jersey Department of Health, Trenton, NJ; Marin County Health and Human Services, San Rafael, CA; Polk County Health Center, Bolivar, MO; Randolph County Public Health, Asheboro, NC; South Dakota State Health Department, Pierre, SD; Springfield-Greene County Health Department, Springfield, MO; Washington State Department of Health, Tumwater, WA; Vermont Department of Health, Burlington, VT; Jordan Moody.

