## Supplemental Digital Content for "Estimates of cases and hospitalizations averted by COVID-19 case investigation and contact tracing in 14 health jurisdictions in the United States"

[**]Contact Tracing Impact Group (alphabetically listed): Greta Anschuetz, Robert Bonacci, Brittany Byers, Joshua Clayton, Nickolas DeLuca, Matthew Donahue, Catherine Donovan, Veronica Fialkowski, Aaron Fleischauer, Heather Forbes, Clay Goddard, Heather Grome, Gibbie Harris, Susan Hayes, Blake Hendrickson, Julia Janssen, Sara Johnson, Amanda Jones, Catherine J. Knott, Reed Magleby, Stephen McCurdy, Alana McGrath, Heather McLaughlin, James Miller, Zack Moore, Michelle Morris, Jill Moses, Allison Newman, Sai Paritala, Caroline Q. Pratt, Lauren Prinzing, Pratima Raghunathan, Jonathan Steinberg, Alana Sulka, Christina G. Tan, Melanie Taylor, Puthiery Va, Kate Varela, Andee Weisbeck, Matthew Willis.

[†]**Corresponding Author:** Gabriel Rainisch

**Author Affiliations:** CDC COVID-19 Response Team

#### Data

We used a previously published evaluation study that assessed the COVID-19 case investigation and contact tracing efforts of 14 health departments (HDs) in 11 states and one tribal nation in the United States over 4-week asynchronous periods during June – October 2020, covering a total of 20 million population.[1] The study focused on assessing process indicators of the completeness and timeliness of case investigation and contact tracing (CICT) program implementation such as the proportions of cases interviewed, contacts notified and monitored, and the time between case identification and contact notification. We obtained the COVID-19 daily incidence data from the Centers for Disease Control and Prevention (CDC)'s COVID Data Tracker.[2] The tribal jurisdiction included in this data set is not represented in the CDC COVID Tracker database, and the data was directly provided to us by the jurisdiction.

#### Epidemiological Inputs

COVIDTracer Advanced[3] is a spreadsheet-based compartmental Susceptible-Exposed-Infectious-Recovered (SEIR) epidemiological model, which illustrates the spread of a pathogen, resultant disease, and impact of interventions in a user-defined population. Readers can download the tool and enter input values of their choosing, exploring the impact of scenarios and assumptions beyond those covered in this manuscript. To model the clinical progression and transmission of disease using COVIDTracer Advanced, we used the following definitions and assumptions. A "case" was defined as a person who has been exposed, infected and subsequently becomes infectious, regardless of the presence of clinical symptoms. We assumed that for the first 3 days after exposure and infection, cases do not infect others. During days 4–5 post-infection, cases are pre-symptomatic but shed virus in amounts that may infect others.[4-7] During days 6–14, the infected person can be symptomatic and shedding virus, albeit during days 11–14 the risk of onward transmission is relatively low but non-zero (the complete infectivity distribution is given in Table A1). We assumed that approximately 40% of cases are asymptomatic during days 6-14 yet have a risk of onward transmission equal to 75% of symptomatic cases (Table A2).

**Table A1**: Daily percentage risk of onward transmission by state of infectiousness and clinical symptoms.

| Day Post exposure- and -infection | Daily percentage of risk of onward transmission[a] | Infectious person's state |
|---|---|---|
| 1 | 0.00% | *Infected, not yet infectious* |
| 2 | 0.00% | |
| 3 | 0.00% | |
| 4 | 16.78% | *Infectious, pre-symptomatic* |
| 5 | 18.03% | |
| 6 | 17.07% | *Infectious, symptomatic* |
| 7 | 14.52% | |
| 8 | 11.27% | |
| 9 | 8.10% | |
| 10 | 5.48% | |
| 11 | 3.55% | |
| 12 | 2.26% | |
| 13 | 1.46% | |
| 14 | 1.48% | |
| **Total** | **100%** | |

[a]Percentages show when onward transmission might occur by day of infectiousness

**Sources:** He *et al.*[4, 5] and Ferretti *et al.*[6]. See also COVIDTracer Advanced manual.[10]

**Table A2**: Epidemiological parameters, values, and sources.

| Parameter | Default Value | Source |
|---|---|---|
| Infected but not yet infectious period | 3 days | CDC COVID-19 Pandemic Planning Scenarios[7] |
| Pre-symptomatic and contagious (infectious) period | 2 days | He *et al*.[4, 5], Ferretti *et al.*[6] |
| Symptomatic and contagious (infectious) period | 9 days | He *et al*.[4, 5], Ferretti *et al.*[6] |
| New infections per case ($R_0$) | 2.5 | CDC COVID-19 Pandemic Planning Scenarios[7] |
| % of cases that are asymptomatic | 40% | CDC COVID-19 Pandemic Planning Scenarios[7] |
| Infectiousness of asymptomatic cases (relative to symptomatic cases) | 75% | CDC COVID-19 Pandemic Planning Scenarios[7] |

**Table A3:** Assumed proportion of cases by age group and infection-to-hospitalization rate, default values in COVIDTracer Advanced and sources.

| Age group (year) | % of Total Cases | Source | % of all cases admitted to hospital care | Source |
|---|---|---|---|---|
| 0 to 17 | 15% | CDC COVID Data Tracker[2] | 0.21 | CDC COVID-19 Response Team[8], Wu *et al.*[9] |
| 18 to 64 | 55% | | 2.17 | |
| 65+ | 30% | | 4.12 | |

**Figure A1 (locations 1-9*)**. Fitted cumulative epidemic curve output from COVIDTracer Advanced and observed data for the 60-day period beginning at the time program evaluations began at each location.

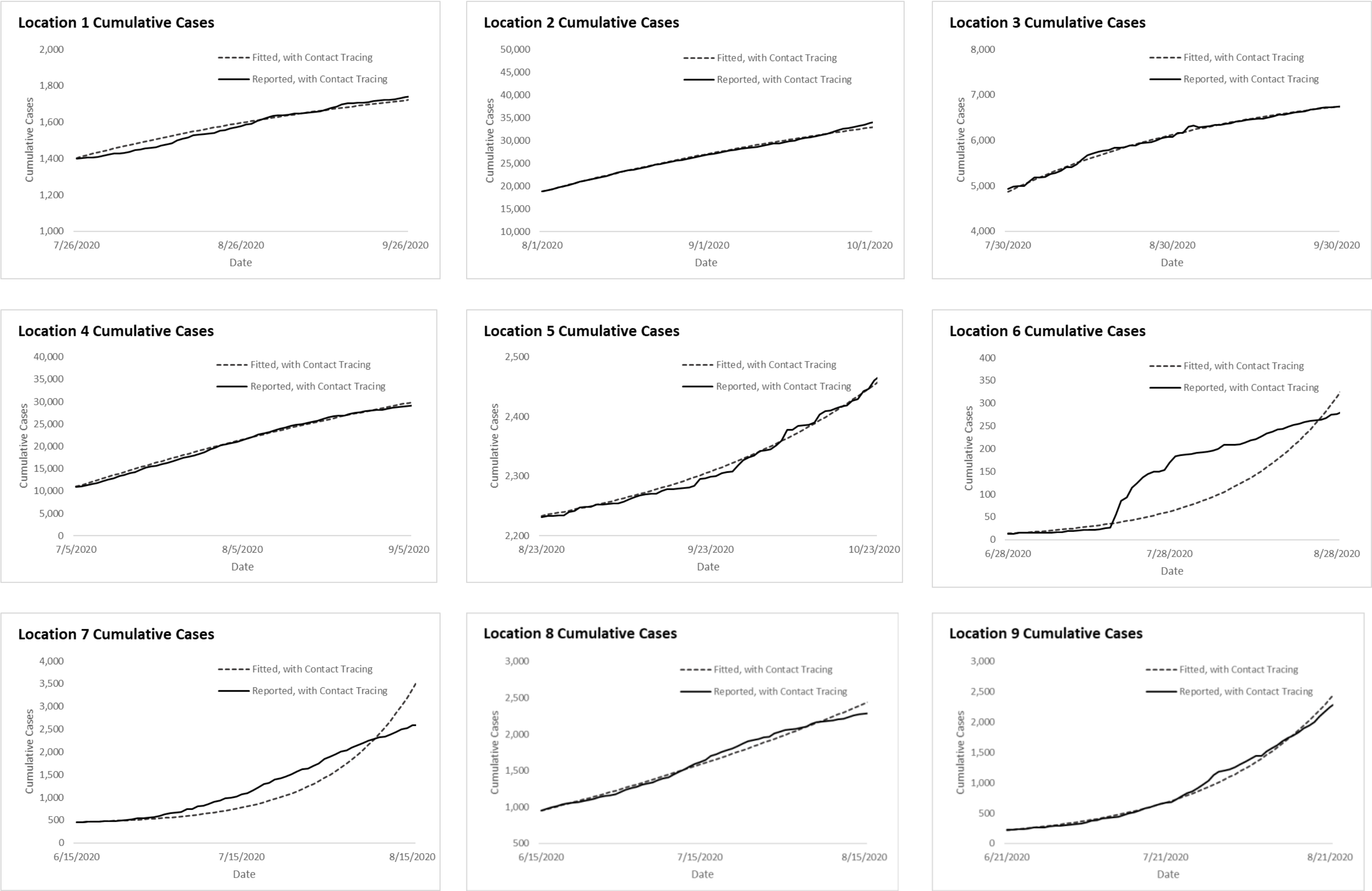


*** Locations 1-4 here correspond to locations 1-4 in the main text.**

**Figure A1 Continued (locations 10-14)**. Fitted cumulative epidemic curve output from COVIDTracer Advanced and observed data for the 60-day period beginning at the time program evaluations began at each location.

**Location 10 not fitted due to unavailable data on days from case identification to isolation**

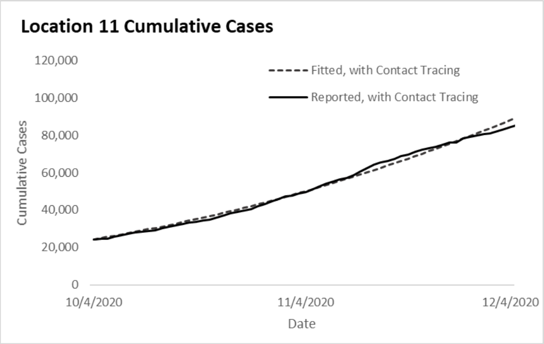


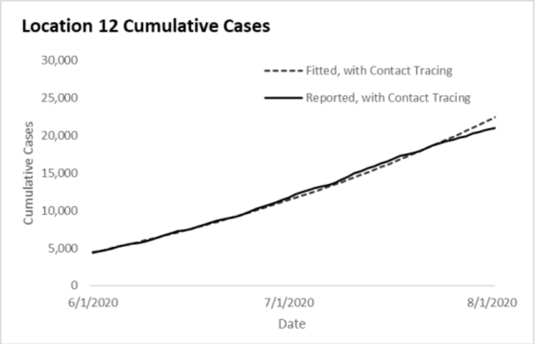


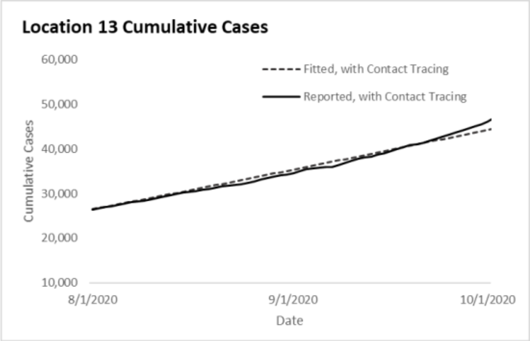


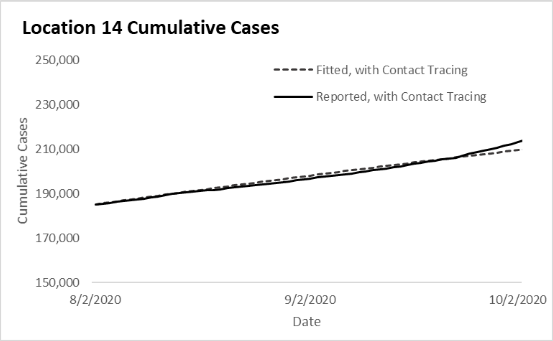

#### Case Investigation and Contact Tracing Effectiveness

The effectiveness of case investigation and contact tracing is determined by the proportion of cases and their contacts that are effectively isolated and quarantined, preventing further transmission in the susceptible population. We assumed that confirmed cases are effectively isolated following case interviews. We further assumed that contacts are quarantined upon either contact notification, or through active monitoring. If infected contacts are left undetected, they will infect additional contacts. Therefore, we calculated the average proportion of cases and contacts isolated and quarantined for each location as follows:

Equation 1:

Contacts are assumed to be quarantined upon notification (hereafter named as "high effectiveness"):

[% Cases interviewed + $R_0$ * % Contacts identified * % Contacts notified)] / (1+$R_0$)

Equation 2:

Contacts are assumed to be quarantined through active monitoring (hereafter named as "low effectiveness"):

[% Cases interviewed + ($R_0$ * % Contacts identified * % Contacts monitored)] / (1+$R_0$)

where $R_0$ is the assumed number of new infections per case without any interventions and when the population is entirely susceptible to infection (Table A2). The % Cases interviewed, % Contacts notified, and % Contacts monitored were process metrics gathered in the evaluation study,[1] and % Contacts identified was an intermediate value calculated as the number of named contacts divided by the expected number of contacts per case:

### Contacts named / (# Cases reported * Average # Contacts per Case in each location)

In general, the % of Contacts notified was higher than the % of Contacts actively monitored. Therefore, the estimated effectiveness was higher when contacts were assumed to be quarantined upon notification. Likewise, the estimated effectiveness was lower when active monitoring was assumed to be required for contact quarantine.

In addition, reducing the time from case identification to effective isolation is critical for case investigation and contact tracing to succeed. The longer the cases and contacts interact with the susceptible population, the greater the opportunity for onward transmission. In practice, cases with no known exposure can be identified and isolated only after symptom onset, and cases with known exposures (*i.e.*, contacts that eventually become infected cases) can begin quarantine upon contact notification (even potentially prior to symptom onset). We assumed asymptomatic cases can only be identified and isolated if they are notified through case investigation and contact tracing.

For the purposes of our study we assumed the proportions of cases with no known exposure and cases with known exposures were equal (*i.e.*, 50/50 breakdown) since we didn't have data on what prompted case identification in each location. Therefore, for each location the days to effective case isolation was determined by taking the average of the days to effective isolation between case groups with known and no known exposures. The time to effective case isolation for each of the two case groups was determined as follows:

For symptomatic cases with no known exposures (*i.e.*, symptoms prompt identification):

We assumed cases experience a 5-day pre-symptomatic period (See Table A2), get tested the next day after symptom onset, wait the number of days observed by Lash *et al.*[1] to learn of their positive result, and begin effective isolation the day after learning this result. See the "Index Case" row in Figure A2 for a visual depiction of this timeline.

For cases with known exposures (*i.e.*, those who were notified they were a contact and eventually became a case):

We assumed contacts quarantine the day after being notified as a contact and that these individuals could contribute to onward transmission based on when they were exposed. Since we did not have information on when exposures actually occurred, we assumed these individuals' exposures occurred at the midpoint of their potential exposure window (in days). We identified the earliest date in this window as the first day of infectiousness among cases to which contacts were exposed. Based on our assumed 5-day pre-symptomatic period for symptomatic cases (described above), this was two days prior to the symptom onset date in cases exposing the contact. We identified the latest possible exposure date as the date the cases exposing them were notified of their positive case status (since they began isolation the next day). See both "Contacts" rows in Figure A2 for a visual depiction of this timeline.

The days between cases with known exposures becoming infected and their exposure notification can vary from what we assumed. For example, cases may take longer to become symptomatic, or get tested the same day they become symptomatic or begin their isolation on the same day as their results notification. Similarly, contacts which become cases may be exposed earlier or later than we assumed and may make up a larger or smaller share of the case pool. Table A6 shows the impact of varying our assumed time to case isolation by 1-day higher and lower prior to completing our fitting procedure for determining the share of transmission reductions attributable to NPIs and Case Investigation and Contact Tracing. The sensitivity analysis associated with 1-day changes described and shown in the main text (Figures 2 and A3) is different in that it shows the impact of varying notification speed when NPI effects are held constant (*i.e.*, given the originally derived share of transmission reductions attributable to NPIs, how speeding or slowing notification would have affected averted cases).

**Figure A2.** Illustrative example of the timing of case isolation and contact quarantine based on reported data from Location 1

| | Day 1 | Day 2 | Day 3 | Day 4 | Day 5 | Day 6 | Day 7 | Day 8 | Day 9 | Day 10 | Days from Exposure to Isolation |
|---|---|---|---|---|---|---|---|---|---|---|---|
| **Index Case** | Exposed | | | Begin Contagious | | Symptom Onset | Tested | | Results Notification | Begin Isolation | 9 |
| **Contacts** (Earliest possible exposure) | | | | Exposed | | | | | Exposure Notification | Begin Quarantine | 6 |
| **Contacts** (Latest possible exposure) | | | | | | | | | Exposed | Begin Quarantine | 1 |

Notes: Location 1 reported 2 days from specimen collection to results notification and 2 days from specimen collection to contact notification (Table 1). The index case (symptomatic case with no known exposure) began showing symptoms on day 6 post exposure-and-infection, got tested on day 7, and was notified of the positive test results on day 9. Its contacts (cases with known exposure) were exposed between day 4 to 9 and notified of their exposure on day 9. Therefore, both the index case and its contacts began isolation and quarantine on day 10.

**FIGURE A3.** Effects of improvements and constraints to case investigation and contact tracing performance measures on the baseline percent cases averted by the program* when monitoring contacts is assumed necessary for effective quarantine (Low CICT effectiveness approach)

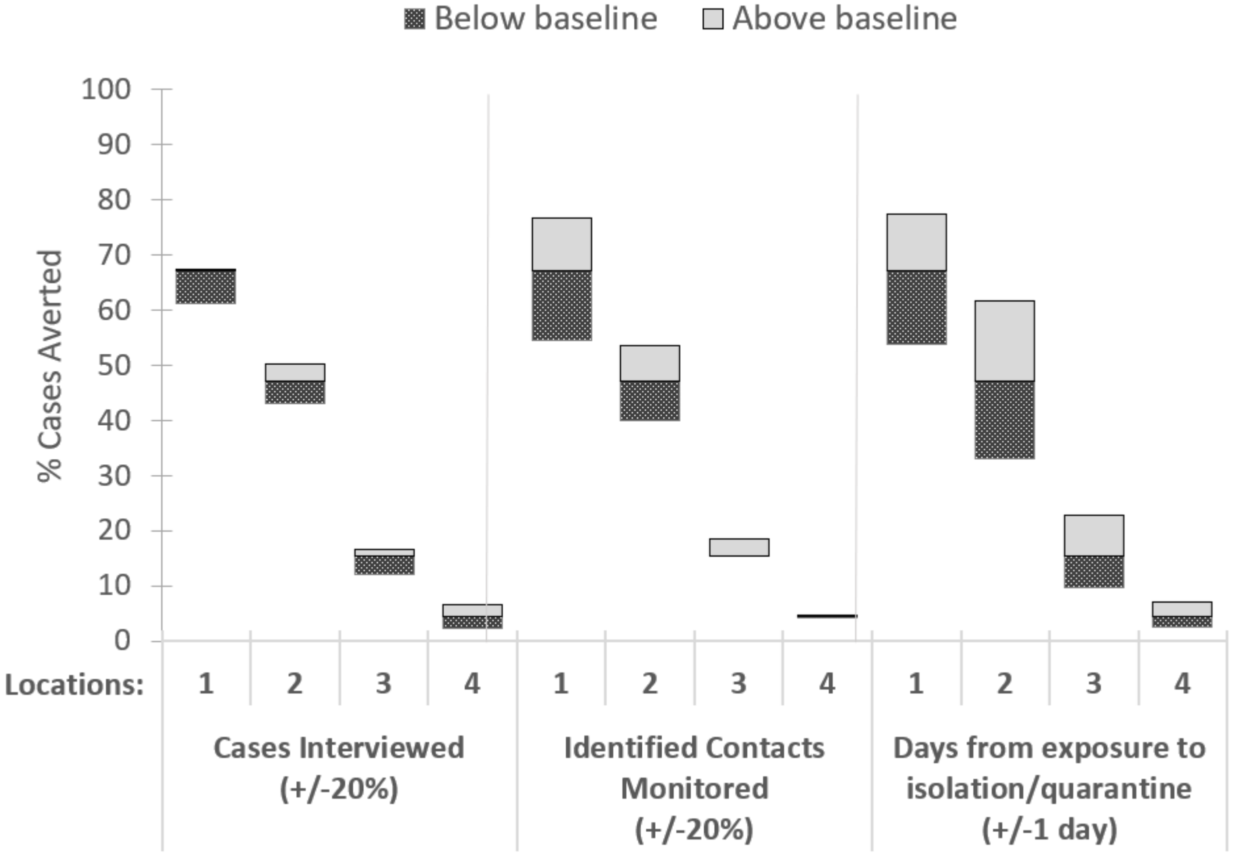


*** Percent cases averted by cases investigated and contact tracing (CICT) calculated as percentage of total cases averted if only nonpharmaceutical interventions (NPIs) were implemented.**

**Notes.** Baseline results shown in Table 2. Cases Interviewed and Contacts Monitored were capped at 100% and 0% when the baseline percentage interviewed was greater than 80% or less than 20%. Similar figure in the main text (Figure 2) is for the High CICT effectiveness scenario: when simply notifying contacts is assumed sufficient to trigger effective quarantine.

**Table A4 (locations 1-7).** Estimated impacts of case investigation and contact tracing (CICT), and nonpharmaceutical interventions (NPIs), by location[a] over 60-day period after contact tracing evaluations initiated

| | **Location 1** | **Location 2** | **Location 3** | **Location 4** | **Location 5** | **Location 6** | **Location 7** |
|---|---|---|---|---|---|---|---|
| Scenario Inputs | | | | | | | |
| **Population** | Category B | Category A | Category C | Category A | Category C | Category C | Category C |
| **Start date of observation** | 7/26/2020 | 8/1/2020 | 7/30/2020 | 7/5/2020 | 8/23/2020 | 6/28/2020 | 6/15/2020 |
| **COVID-19 cases** | | | | | | | |
| Total cases before the start of observation | 1 396 | 18 488 | 4 817 | 10 724 | 2 231 | 13 | 453 |
| Cases in the last 14 days | 113 | 4 686 | 870 | 3 853 | 43 | 5 | 33 |
| Trend in the last 14 days | plateaued | plateaued | plateaued | slowly increasing | slowly decreasing | slowly increasing | plateaued |
| **CICT Effectiveness** | | | | | | | |
| % of cases and contacts isolated[b] | 39.5 – 86.0% | 52.7 – 55.0% | 26.1 – 50.5% | 11.8 – 12.1% | 76.6 – 81.4% | 70.3 – 71.0% | 21.4 – 30.2% |
| Days from infection to isolation[c] | 6 days | 8 days | 9 days | 10 days | 6 days | 6 days | 7 days |
| Scenario Results | | | | | | | |
| **Transmission Fraction** | | | | | | | |
| Reduction from CICT | 8.6 – 26.2% | 5.0 – 5.2% | 1.4 – 2.7% | 0.4 – 0.4% | 28.7 – 31.7% | 27.8 – 28.1% | 4.8 – 7.0% |
| Reduction from NPIs[d] | 54.6 – 36.6% | 57.6 – 57.3% | 63.5– 62.0% | 61.0 – 61.0% | 22.2 – 19.1% | 18.0 – 17.7% | 32.8 – 30.9% |
| Remaining Transmission (100% minus above values) | 36.8 – 37.2% | 37.4 – 37.5% | 35.1 – 35.3% | 38.6 – 38.6% | 49.1 – 49.2% | 54.2 – 54.2% | 62.4 – 62.1% |
| **Additional Cases Averted by CICT (%), 60 days[e]** | 651 – 9 480 (67.1 – 96.8%) | 12 598 – 13 568 (47.1 – 48.8%) | 344 – 768 (15.4 – 28.8%) | 859 – 882 (4.4 – 4.5%) | 5 238 – 6 879 (96.1 – 97.0%) | 4 319 – 4 450 (93.8 – 94.0%) | 1 684 – 2 738 (38.3 – 50.0%) |
| **Additional Hospitalizations Averted by CICT (%), 60 days[e]** | 16 – 233 (67.1 – 96.8%) | 310 – 333 (47.1 – 48.8%) | 8 – 19 (15.4 – 28.8%) | 21 – 22 (4.4 – 4.5%) | 129 – 160 (96.1 – 97.0%) | 106 – 109 (93.8 – 94.0%) | 41 – 67 (38.3 – 50.0%) |

† To preserve anonymity, populations are categorized by population size as follows: Category A > 1 million; Category B >500 000 to ≤ 1 million; Category C: ≤ 500 000.

[a]Case investigation and contact tracing implemented per scenarios in Table 4A and effects were assumed constant over 60 days.

[b]Calculated as follows using values observed at locations during case investigation and contact tracing evaluations and an assumed $R_0$=2.5:

Minimum (low) value assumes monitoring is required for effective quarantine of contacts:

[% Cases interviewed + ($R_0$ * % Contacts identified * % Contacts actively monitored)] /(1+$R_0$)

Maximum (high) value assumes contact notification is sufficient for effective quarantine of contacts:

[% Cases interviewed + ($R_0$ * % Contacts identified * % Contacts notified)] / (1+$R_0$)

[c]The average length of time from exposure-and-infection to isolation (including contacts which later became cases). See CICT Effectiveness in the Supplement.

[d] NPI interventions including masks use, social distancing, school and restaurant closures, etc. Low NPI effectiveness values were generated with the fitting process when CICT effectiveness was high; similarly, high NPI effectiveness values were generated when CICT effectiveness was low.

[e] Percent calculated as (Total Cases Averted when both CICT and NPIs implemented - Total Cases Averted when only NPIs implemented) / Total Cases Averted when both CICT and NPIs implemented). For example, for every 100 cases prevented by nonpharmaceutical interventions in locations 1-4, CICT averted between 4.4 and 96.8 additional cases.

**TABLE A4 Continued (locations 8-14).** Estimated impacts of case investigation and contact tracing (CICT), and nonpharmaceutical interventions (NPIs), by location[a] over 60-day period after contact tracing evaluations initiated

| | Location 8 | Location 9 | Location 10[f] | Location 11 | Location 12 | Location 13 | Location 14 |
|---|---|---|---|---|---|---|---|
| Scenario Inputs | | | | | | | |
| **Population** | Category C | Category C | Category B | Category B | Category A | Category A | Category A |
| **Start date of observation** | 6/15/2020 | 6/21/2020 | 7/1/2020 | 10/4/2020 | 6/1/2020 | 8/1/2020 | 8/2/2020 |
| **COVID-19 cases** | | | | | | | |
| Total cases before the start of observation | 931 | 218 | 9 237 | 23 988 | 4 307 | 26 211 | 184 358 |
| Cases in the last 14 days | 249 | 44 | 2 221 | 5 545 | 1 685 | 3 850 | 5 835 |
| Trend in the last 14 days | plateaued | rapidly increasing | plateaued | slowly increasing | slowly increasing | plateaued | plateaued |
| **CICT Effectiveness** | | | | | | | |
| % of cases and contacts isolated[b] | 50.6–52.7% | 54.5 – 61.1 | 22.1 – 22.1% | 21.3 – 30.8% | 43.4 – 43.9% | 22.2 – 52.7% | 32.2 – 35.6% |
| Days from infection to isolation[c] | 9 days | 7 days | Not Available | 7 days | 8 days | 8 days | 8 days |
| Scenario Results | | | | | | | |
| **Transmission Fraction** | | | | | | | |
| Reduction from CICT | 3.3 – 3.5% | 11.7 – 13.4% | | 3.6 – 5.4% | 4.7 – 4.8% | 2.2 – 5.5% | 3.1 – 3.5% |
| Reduction from NPIs[d] | 54.0–53.9% | 36.2 – 34.8% | | 49.6 – 47.9% | 51.2 – 51.1% | 56.7 – 53.4% | 56.3 – 55.9% |
| Remaining Transmission (100% minus above values) | 42.7–42.6% | 52.1 – 51.8% | | 46.8 – 46.7% | 44.1 – 44.1% | 41.1 – 41.1% | 40.6 – 40.6% |
| **Additional Cases Averted by CICT (%), 60 days[e]** | 733 – 768 (33.3–34.5%) | 4 857–6 112 (70.0–74.7%) | | 28 141–45 869 (31.0–42.2%) | 12 608–12 904 (42.0 – 42.4%) | 5 386–17 264 (23.2–49.3%) | 11 634–13 378 (32.3 – 35.3%) |
| **Additional Hospitalizations Averted by CICT (%), 60 days[e]** | 18 – 19 (33.3–34.5%) | 119–150 (70.0–74.7%) | | 692–1 127 (31.0–42.2%) | 310 – 317 (42.0 – 42.4%) | 132–424 (23.2–49.3%) | 286 – 329 (32.3 – 35.3%) |

† To preserve anonymity, populations are categorized by population size as follows: Category A > 1 million; Category B >500 000 to ≤ 1 million; Category C: ≤ 500 000.

[a]Case investigation and contact tracing implemented per scenarios in Table 4A and effects were assumed constant over 60 days.

[b]Calculated as follows using values observed at locations during case investigation and contact tracing evaluations and an assumed $R_0$=2.5:

Minimum (low) value assumes monitoring is required for effective quarantine of contacts:

[% Cases interviewed + ($R_0$ * % Contacts identified * % Contacts actively monitored)] /(1+$R_0$)

Maximum (high) value assumes contact notification is sufficient for effective quarantine of contacts:

[% Cases interviewed + ($R_0$ * % Contacts identified * % Contacts notified)] / (1+$R_0$)

[c]The average length of time from exposure-and-infection to isolation (including contacts which later became cases). See CICT Effectiveness in the Supplement.

[d] NPI interventions including masks use, social distancing, school and restaurant closures, etc. Low NPI effectiveness values were generated with the fitting process when CICT effectiveness was high; similarly, high NPI effectiveness values were generated when CICT effectiveness was low.

[e] Percent calculated as (Total Cases Averted when both CICT and NPIs implemented - Total Cases Averted when only NPIs implemented) / Total Cases Averted when both CICT and NPIs implemented). For example, for every 100 cases prevented by nonpharmaceutical interventions in locations 1-4, CICT averted between 4.4 and 96.8 additional cases.

[f] Location 10 did not report the median days from case identification to isolation, thus the impact of CICT could not be estimated.

**TABLE A5(a).** Estimated cases averted with improving and constraining case investigation and contact tracing performance measures relative to baseline, assuming contact notification is a sufficient trigger for contacts to effectively quarantine, by location, over 60-days after program evaluations were initiated

| Performance measure | **Location 1**<br>Baseline Values:<br>Cases Interviewed = 99%<br>Contacts Notified = 95%<br>Days to isolation = 6 | **Location 2**<br>Baseline Values:<br>Cases Interviewed = 83%<br>Contacts Notified = 61%<br>Days to isolation = 8 | **Location 3**<br>Baseline Values:<br>Cases Interviewed = 91%<br>Contacts Notified = 85%<br>Days to isolation = 9 | **Location 4**<br>Baseline Values:<br>Cases Interviewed = 33%<br>Contacts Notified = 54%<br>Days to isolation = 10 |
|---|---|---|---|---|
| **Cases Interviewed (%)** | | | | |
| Baseline +20%[a] | 9 486 (96.8%) | 14 431 (52.0%) | 856 (32.1%) | 1 286 (6.6%) |
| Baseline | 9 480 (96.8%) | 13 568 (48.8%) | 768 (28.8%) | 882 (4.5%) |
| Baseline -20% | 9 386 (95.8%) | 12 509 (45.0%) | 692 (26.0%) | 471 (2.4%) |
| **Contacts Notified (%)** | | | | |
| Baseline +20%[a] | 9 525 (97.2%) | 15 314 (55.1%) | 824 (30.9%) | 953 (4.9%) |
| Baseline | 9 480 (96.8%) | 13 568 (48.8%) | 768 (28.8%) | 882 (4.5%) |
| Baseline -20% | 9 245 (94.4%) | 11 605 (41.8%) | 691 (25.9%) | 810 (4.2%) |
| **Median days from cases being infected to their contacts isolated** | | | | |
| 3 days faster than Baseline | --[b] | 23 479 (84.5%) | 1 775 (66.6%) | 3 289 (16.9%) |
| 2 days faster than Baseline | --[b] | 21 106 (76.0%) | 1 451 (54.5%) | 2 232 (11.5%) |
| 1 day faster than Baseline | 9 703 (99.9%) | 17 667 (63.6%) | 1 096 (41.1%) | 1 435 (7.4%) |
| Baseline | 9 480 (96.8%) | 13 568 (48.8%) | 768 (28.8%) | 882 (4.5%) |
| 1 day slower than Baseline | 8 753 (89.3%) | 9 572 (34.5%) | 504 (18.9%) | 520 (2.7%) |
| 2 days slower than Baseline | 7 269 (74.2%) | 6 281 (22.6%) | 312 (11.7%) | 291 (1.5%) |
| 3 days slower than Baseline | 5 343 (54.5%) | 3 879 (14.0%) | 181 (6.8%) | 145 (0.7%) |

**Notes**

[a]This value was capped at 100% when the baseline percentage interviewed was greater than 80%.

[b]This value not calculated since the minimum median days from case infection to contacts being isolated is six (5 days from infection to symptom onset + 1 day for testing).

**TABLE A5(b).** Estimated cases averted with improving and constraining case investigation and contact tracing performance measures relative to baseline, assuming active monitoring is required for effective contact quarantine, by location, over 60-days after program evaluations were initiated

| Performance measure | Location 1<br>Baseline Values:<br>Cases Interviewed = 99%<br>Contacts Monitored = 19%<br>Days to isolation = 6 | Location 2<br>Baseline Values:<br>Cases Interviewed = 83%<br>Contacts Monitored = 56%<br>Days to isolation = 8 | Location 3<br>Baseline Values:<br>Cases Interviewed = 91%<br>Contacts Monitored = 0%<br>Days to isolation = 9 | Location 4<br>Baseline Values:<br>Cases Interviewed = 33%<br>Contacts Monitored = 48%<br>Days to isolation = 10 |
|---|---|---|---|---|
| **Cases Interviewed (%)** | | | | |
| Baseline +20%[a] | 654 (67.5%) | 13 448 (50.3%) | 375 (16.7%) | 1 263 (6.5%) |
| Baseline | 651 (67.1%) | 12 598 (47.1%) | 344 (15.4%) | 859 (4.4%) |
| Baseline -20% | 594 (61.3%) | 11 555 (43.2%) | 273 (12.2%) | 448 (2.3%) |
| **Contacts Quarantined (%)** | | | | |
| Baseline +20%[a] | 744 (76.8%) | 14 319 (53.5%) | 414 (18.5%) | 931 (4.8%) |
| Baseline | 651 (67.1%) | 12 598 (47.1%) | 344 (15.4%) | 859 (4.4%) |
| Baseline -20%[b] | 529 (54.6%) | 10 664 (39.9%) | 344 (15.4%) | 787 (4.1%) |
| **Median days from cases being infected to their contacts isolated** | | | | |
| 3 days faster than Baseline | --‡ | 22 207 (83.0%) | 934 (41.7%) | 3 209 (16.5%) |
| 2 days faster than Baseline | --‡ | 19 843 (74.2%) | 715 (31.9%) | 2 177 (11.2%) |
| 1 day faster than Baseline | 750 (77.4%) | 16 499 (61.7%) | 512 (22.8%) | 1 399 (7.2%) |
| Baseline | 651 (67.1%) | 12 598 (47.1%) | 344 (15.4%) | 859 (4.4%) |
| 1 day slower than Baseline | 522 (53.9%) | 8 851 (33.1%) | 220 (9.8%) | 507 (2.6%) |
| 2 days slower than Baseline | 386 (39.8%) | 5 794 (21.7%) | 135 (6.0%) | 283 (1.5%) |
| 3 days slower than Baseline | 264 (27.2%) | 3 573 (13.4%) | 77 (3.5%) | 141 (0.7%) |

**Notes**

[a]This value was capped at 100% when the baseline percentage interviewed was greater than 80%.

[b]This value was kept at 0% when the baseline percentage monitored was less than 20%.

**TABLE A6(a).** Estimated impacts of case investigation and contact tracing and nonpharmaceutical interventions in Location 1 over 60-day period after program evaluations were initiated, varying the days from infection to isolation

| **Days from Infection to Isolation** | **5** | **6 (default)** | **7** |
|---|---|---|---|
| **Transmission Fraction** | | | |
| Reduction from CICT | 12.8 – 47.5% | 8.6 – 26.2% | 5.6 – 15.1% |
| Reduction from All Other Interventions (NPIs) | 50.5 – 15.2% | 54.6 – 36.6% | 57.6 – 47.9% |
| Remaining Transmission (100% minus above values) | 36.7 – 37.3% | 36.8 – 37.2% | 36.8 – 37.0% |
| **Cases Averted by CICT (%), 60 days** | 1 338 – 97 014 (80.8 – 99.7%) | 651 – 9 480 (67.1 – 96.8%) | 339 – 2 009 (51.4 – 86.3%) |
| **Hospitalizations Averted by CICT (%), 60 days** | 33 – 2 433 (80.8 – 99.7%) | 16 – 233 (67.1 – 96.8%) | 8 – 49 (51.4 – 86.3%) |

**TABLE A6(b).** Estimated impacts of case investigation and contact tracing and nonpharmaceutical interventions in Location 2 over 60-day period after program evaluations were initiated, varying the days from infection to isolation

| **Days from Infection to Isolation** | **7** | **8 (default)** | **9** |
|---|---|---|---|
| **Transmission Fraction** | | | |
| Reduction from CICT | 8.1 – 8.5% | 5.0 – 5.2% | 3.0 – 3.2% |
| Reduction from All Other Interventions (NPIs) | 54.5 – 54.0% | 57.6 – 57.3% | 59.6 – 59.4% |
| Remaining Transmission (100% minus above values) | 37.4 – 37.5% | 37.4 – 37.5% | 37.4 – 37.4% |
| **Cases Averted by CICT (%), 60 days** | 25 552 – 28 102 (64.4 – 66.4%) | 12 598 – 13 568 (47.1 – 48.8%) | 6 663 – 7 108 (32.0 – 33.3%) |
| **Hospitalizations Averted by CICT (%), 60 days** | 628 – 691 (64.4 – 66.4%) | 310 – 333 (47.1 – 48.8%) | 164 – 175 (32.0 – 33.3%) |

**TABLE A6(c).** Estimated impacts of case investigation and contact tracing and nonpharmaceutical interventions in Location 3 over 60-day period after program evaluations were initiated, varying the days from infection to isolation

| **Days from Infection to Isolation** | **8** | **9 (default)** | **10** |
|---|---|---|---|
| **Transmission Fraction** | | | |
| Reduction from CICT | 2.2 – 4.5% | 1.4 – 2.7% | 0.8 – 1.6% |
| Reduction from All Other Interventions (NPIs) | 62.7 – 60.2% | 63.6– 62.0% | 64.1 – 63.2% |
| Remaining Transmission (100% minus above values) | 35.1 – 35.3% | 35.1 – 35.3% | 35.1 – 35.2% |
| **Cases Averted by CICT (%), 60 days** | 573 – 1 407 (23.3 – 42.7%) | 344 – 768 (15.4 – 28.8%) | 203 – 427 (9.7 – 18.4%) |
| **Hospitalizations Averted by CICT (%), 60 days** | 14 – 35 (23.3 – 42.7%) | 8 – 19 (15.4 – 28.8%) | 5 – 10 (9.7 – 18.4%) |

**TABLE A6(d).** Estimated impacts of case investigation and contact tracing and nonpharmaceutical interventions in Location 4 over 60-day period after program evaluations were initiated, varying the days from infection to isolation

| **Days from Infection to Isolation** | **9** | **10 (default)** | **11** |
|---|---|---|---|
| **Transmission Fraction** | | | |
| Reduction from CICT | 0.7 – 0.7% | 0.4 – 0.4% | 0.2 – 0.2% |
| Reduction from All Other Interventions (NPIs) | 60.8 – 60.8% | 61.0 – 61.0% | 61.2 – 61.2% |
| Remaining Transmission (100% minus above values) | 38.5 – 38.5% | 38.6 – 38.6% | 38.6 – 38.6% |
| **Cases Averted by CICT (%), 60 days** | 1 438 – 1 476 (7.2 – 7.4%) | 859 – 882 (4.4 – 4.5%) | 493 – 506 (2.6 – 2.7%) |
| **Hospitalizations Averted by CICT (%), 60 days** | 35 – 36 (7.2 – 7.4%) | 21 – 22 (4.4 – 4.5%) | 12 – 12 (2.6 – 2.7%) |

**Adjustment for Under-reported and Under-detected Cases**

**Table A7(a).** Estimated impacts of case investigation and contact tracing and nonpharmaceutical interventions over 60-day period after program evaluations were initiated, with and without adjustment for under-reported asymptomatic cases; Locations 1 and 2

| | Location 1<br>Baseline | Location 1<br>Multiplying reported cases by x2 | Location 2<br>Baseline | Location 2<br>Multiplying reported cases by x2 |
|---|---|---|---|---|
| **Transmission Fraction** | | | | |
| Reduction from CICT | 8.6 – 26.2% | 8.7 – 26.3% | 5.0 – 5.2% | 5.0 – 5.3% |
| Reduction from All Other Interventions (NPIs) | 54.6 – 36.6% | 54.5 – 36.4% | 57.6 – 57.3% | 57.3 – 57.1% |
| Remaining Transmission (100% minus above values) | 36.8 – 37.2% | 36.8 – 37.3% | 37.4 – 37.5% | 37.7 – 37.6% |
| **Cases Averted by CICT (%), 60 days** | 651 – 9 480<br>(67.1 – 96.8%) | 1 296 – 18 707<br>(67.0 – 96.7%) | 12 598 – 13 568<br>(47.1 – 48.8%) | 25 173 – 26 719<br>(46.9 – 48.6%) |
| **Hospitalizations Averted by CICT (%), 60 days** | 16 – 233<br>(67.1 – 96.8%) | 32 – 460<br>(67.0 – 96.7%) | 310 – 333<br>(47.1 – 48.8%) | 619 – 657<br>(46.9 – 48.6%) |

**Table A7(b).** Estimated impacts of case investigation and contact tracing and nonpharmaceutical interventions over 60-day period after program evaluations were initiated, with and without adjustment for under-reported asymptomatic cases; Locations 3 and 4

| | Location 3<br>Baseline | Location 3<br>Multiplying reported cases by x2 | Location 4<br>Baseline | Location 4<br>Multiplying reported cases by x2 |
|---|---|---|---|---|
| **Transmission Fraction** | | | | |
| Reduction from CICT | 1.4 – 2.7% | 1.4 – 2.8% | 0.4 – 0.4% | 0.4 – 0.4% |
| Reduction from All Other Interventions (NPIs) | 63.5 – 62.0% | 62.7 – 61.2% | 61.0 – 61.0% | 60.4 – 60.4% |
| Remaining Transmission (100% minus above values) | 35.1 – 35.3% | 35.9 – 36.0% | 38.6– 38.6% | 39.2 – 39.2% |
| **Cases Averted by CICT (%), 60 days** | 344 – 768<br>(15.4 – 28.8%) | 678 – 1 501<br>(15.2 – 28.5%) | 859 – 882<br>(4.4 – 4.5%) | 1 674 – 1 718<br>(4.3 – 4.4%) |
| **Hospitalizations Averted by CICT (%), 60 days** | 8 – 19<br>(15.4 – 28.8%) | 17 – 37<br>(15.2 – 28.5%) | 21 – 22<br>(4.4 – 4.5%) | 41 – 42<br>(4.3 – 4.4%) |

2. CDC COVID Data Tracker. https://covid.cdc.gov/covid-data-tracker/
